# Machine Learning Interpretability Methods to Characterize the Importance of Hematologic Biomarkers in Prognosticating Patients with Suspected Infection

**DOI:** 10.1101/2023.05.30.23290757

**Authors:** Dipak P. Upadhyaya, Yasir Tarabichi, Katrina Prantzalos, Salman Ayub, David C Kaelber, Satya S. Sahoo

## Abstract

Early detection of sepsis in patients admitted to the emergency department (ED) is an important clinical objective as early identification and treatment can help reduce morbidity and mortality rate of 20% or higher. Hematologic changes during sepsis-associated organ dysfunction are well established and a new biomarker called Monocyte Distribution Width (MDW) has been recently approved by the US Food and Drug Administration for sepsis. However, MDW, which quantifies monocyte activation in sepsis patients, is not a routinely reported parameter and it requires specialized proprietary laboratory equipment. Further, the relative importance of MDW as compared to other routinely available hematologic parameters and vital signs has not been studied, which makes it difficult for resource constrained hospital systems to make informed decisions in this regard. To address this issue, we analyzed data from a cohort of ED patients (n=10,229) admitted to a large regional safety-net hospital in Cleveland, Ohio with suspected infection who later developed poor outcomes associated with sepsis. We developed a new analytical framework consisting of seven data models and an ensemble of high accuracy machine learning (ML) algorithms (accuracy values ranging from 0.83 to 0.90) for the prediction of outcomes more common in sepsis than uncomplicated infection (3-day intensive care unit stay or death). To characterize the contributions of individual hematologic parameters, we applied the Local Interpretable Model-Agnostic Explanation (LIME) and Shapley Additive Value (SHAP) interpretability methods to the high accuracy ML algorithms. The ML interpretability results were consistent in their findings that the value of MDW is grossly attenuated in the presence of other routinely reported hematologic parameters and vital signs data. Further, this study for the first time shows that complete blood count with differential (CBC-DIFF) together with vital signs data can be used as a substitute for MDW in high accuracy ML algorithms to screen for poor outcomes associated with sepsis.

## 1. Introduction

Sepsis is a dysregulated host response to infection, and it is a common cause of organ dysfunction, shock, and death. The incidence of sepsis in hospitalized patients is over 6%, with an associated mortality rate of 20% or higher [1, 2]. The Surviving Sepsis Campaign (SSC) has developed several evidence-based guidelines to address the challenge of sepsis detection with 93 recommendations, including the use of sepsis screening tools such as systemic inflammatory response syndrome (SIRS) criteria and the modified early warning score (MEWS) [2]. Several studies have analyzed physiological and laboratory data among other patient information from Electronic Health Record (EHR) systems to compute sepsis risk scores, while other studies have focused on identifying specific biomarkers to aid in the early detection of sepsis [3–7].

These biomarker studies have analyzed the value of widely available laboratory results, such as C-reactive protein (CRP), procalcitonin, and more recently, monocyte distribution width (MDW) in identifying patients with sepsis. Hematologic changes in sepsis-associated organ dysfunction are well established [8, 9]. Various perturbations in routinely measured cell population data have been associated with either early sepsis or have been shown to be prognostic in the context of sepsis [10]. Cell population counts, including white blood cells (leukocytosis or leukopenia), neutrophil, and eosinophil counts have been particularly well studied in this context. In addition to cell counts, more advanced hematologic analyzers are capable of measuring cell size and other parameters from blood cells. Two measures derived from these advanced instruments are mean platelet volume and MDW, both of which have been shown to be strong prognosticators in patients with sepsis [11–13].

As a cytometric parameter that reflects monocyte activation, MDW can be measured by proprietary hematology analyzers as part of otherwise routinely obtained complete blood count with differential (CBC-DIFF) panel [14, 15]. Maximizing the inference of prognostic data from cell population data is valuable primarily due to the high prevalence of blood count data that is obtained in routine practice [16]. MDW has been approved by the US Food and Drug Association (FDA) as marker for the early detection of sepsis in adults admitted to the emergency department (ED). An elevated MDW value of 20 or higher in adult patients as measured by a proprietary cellular analysis system developed by Beckman Coulter, Inc. (Brea, CA, USA) has been associated with an increased risk of developing sepsis within 12 hours of hospital admission [17, 18]. Recent studies have also explored the role of MDW as a prognostic marker in patients with SARS-CoV-2-associated disease (COVID-19) infection [19, 20].

### Relative importance of MDW in screening for sepsis

While the value of MDW as an independent prognosticator is evident, it’s relative value in the detection of clinically significant sepsis is less clear especially in the presence of other readily available hematologic and vital signs data. We are not aware of any existing study that has systematically characterized the role of hematologic parameters, including MDW, to accurately screen for patients at an increased risk of more severe sepsis related outcomes including prolonged intensive care unit (ICU) stay or death. Understanding the relative value of MDW in the detection of sepsis is critical, as not all hematologic analyzers are capable of measuring MDW values, and equipment upgrades or replacements are necessary for healthcare systems interested in leveraging this new biomarker. This objective is important both as a clinical goal and to enable informed decision making for allocation of limited healthcare resources with significant impact on patient outcomes.

### Machine learning algorithms for sepsis screening

Machine learning (ML) algorithms are being used successfully in a variety of medical applications, including their use for sepsis screening [21–23]. Recent studies have used ML algorithms for sepsis screening in both in-patient as well as in ED settings using retrospective multi-modal data, including vital parameters, and text data from the EHR or clinical notes [21, 24]. These previous studies have demonstrated the effectiveness of ML algorithms in the prediction of poor outcomes associated with sepsis with intervention [25], early diagnosis of sepsis using unstructured text from ED notes [26], and for predicting patient survival, as well as length of hospital stay [27]. ML algorithms have also shown higher sensitivity and specificity in sepsis screening as compared to traditional tools such as SIRS, MEWS, and Sequential Organ Failure Assessment (SOFA) [3, 4, 21].

However, there are no existing studies that have used ML algorithms to characterize the role of hematologic biomarkers and specifically MDW in sepsis detection. Our study addresses this critical gap by developing a novel approach that combines an ensemble of high accuracy ML algorithms together with two ML interpretability methods to provide new insights into the role of hematologic parameters in screening for severe sepsis-related outcomes (Figure 1 shows an overview of the study). In the following sections, we describe the method used in this study (Section 2), the results of our study (Section 3), and discuss the significance of these results in the context of using ML interpretability methods for the prognostication of severe sepsis-related outcomes (Section 4).

**Figure 1:**
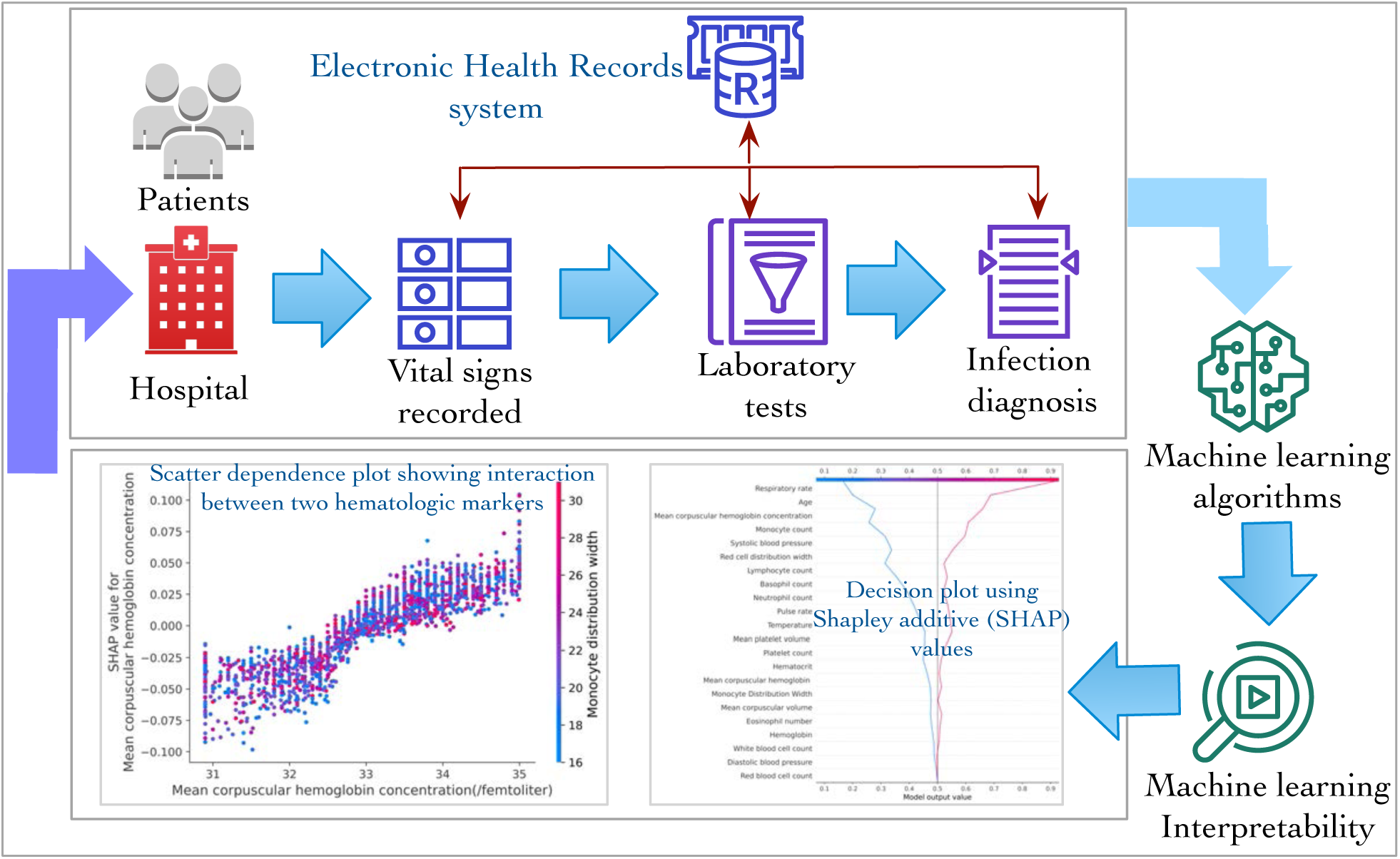
The study analyzes 22 data elements from 10,229 patients admitted to ED at the MetroHealth hospital system to characterize the role of individual hematologic parameters, including MDW, and vital signs in screening for severe sepsis. The data elements were extracted from an EHR system that stores records of 140,000 ED visits per year at the MetroHealth hospital system.

## 2. Methods

### 2.1. Study design, setting, and cohort selection

This retrospective study was reviewed and approved by the MetroHealth hospital system institutional review board (IRB) (approval: STUDY00000097).

#### Study design and data

MetroHealth hospital is a safety-net public health care system in Cleveland, Ohio serving a large metropolitan area of 1.2 million persons. The system averages over 140,000 ED visits per year. All encounters for adult patients presenting to the ED between January 1^st^ 2019 and December 24^th^ 2023, were eligible for inclusion in this study. Encounters that did not have complete cell count and differential data or vital signs data in the ED setting were excluded. In our system, MDW is automatically measured and reported in all cell count with differential tests for adult patients in the ED as required by the FDA guidelines. Encounters for patients that did not meet clinical criteria for infection were excluded. In this study, infection and severe sepsis-related outcomes were defined according to the standards set forth by Seymour et al [28]. Infection (as suspected by clinicians) was defined as the administration of a single dose of an antibiotic followed by the ascertainment of a body fluid culture within 24 hours, or a culture followed by an antibiotic administration within 72 hours. We only included infection when time zero occurred within 72 hours of arrival at the ED and severe sepsis-related outcomes was determined by infection, followed by at least a 3-day intensive care unit stay or death. For our primary outcomes, we leveraged a prolonged (3-day) ICU stay or death as robust clinical outcomes that were more commonly encountered in severe sepsis-related outcomes than in uncomplicated infection. This characterization was based on the third international definition consensus of sepsis (Sepsis-3) identifying qSOFA as a screening mechanism to identify patients with suspected infection who were prone to more severe clinical outcomes characteristic of sepsis. We adopted as similar framework using vital signs and lab markers are predictors of a poor outcome characteristic of sepsis (i.e. a phenotype of severe sepsis). The limitations of this approach mirror the limitations of qSOFA, including high specificity with low sensitivity, and an emphasis on prognostication rather than screening [29].

The patient data was obtained by querying for discrete data from the Epic EHR system (Verona, WI). Patient age was calculated based on the date of arrival (Figure S1 in the supplementary material shows the workflow used to select the study cohort). During the study period, there were 613,341 adult ED visits and out of these visits a total of 67,475 encounters met the study inclusion criteria of having both vital signs and complete hematologic parameter data. From these 67,475 encounters, 10,229 unique patients met the Sepsis-3 criteria for this study. 3484 patients (34%) in the cohort experienced a 3-day intensive care unit stay or death, which led to their classification in the severe sepsis category. Among these patients with severe sepsis-related outcomes,1973 patients stayed in the ICU for more than 3 days and 1511 patients died. Table 1 lists the details of the patient cohort used in this study.

**Table 1:** The distribution of demography, hematologic parameters, and vital signs data of the patients in the study cohort.

| **Data elements** | **Patients with infection**<br>**(n=6745), mean (±SD)** | **Patients with Severe**<br>**Sepsis-related Outcomes**<br>**(n= 3484), mean (±SD)** |
| --- | --- | --- |
| Age (years), mean (SD) | 57.14 (±16.55) | 60.14 (±16.24) |
| **Sex** |  |  |
| Male | 3552 | 2111 |
| Female | 3198 | 1426 |
| **Race** |  |  |
| White | 3865 | 2221 |
| African American | 2247 | 1038 |
| Asian | 61 | 31 |
| American Indian/Native Hawaiian | 71 | 19 |
| Unavailable | 501 | 175 |
| **Vital Signs** |  |  |
| Temperature (Fahrenheit) | 99.11 (±0.59) | 99.48.46 (±0.78) |
| Pulse rate | 94.82 (± 7.98) | 97.93 (±6.43) |
| Respiratory rate | 24.34 (± 2.47) | 27.50 (±5.63) |
| Blood pressure: Systolic/Diastolic (mmHg) | 125.78 (±20.8)/ 71.97 (±14.43) | 105.65 (±26.64)/ 68.4 (±16.54) |
| **Hematologic parameters** |  |  |
| Hematocrit | 39.44. (± 7.17) | 45.23 (±8.81) |
| Hemoglobin | 14.28 (± 1.01) | 14.47 (±0.74) |
| Mean corpuscular hemoglobin (MCH) | 31.42 (± 4.21) | 36.24 (±6.64) |
| Mean corpuscular hemoglobin concentration (MCHC) | 39.32 (±2.1) | 41.54 (±3.34) |
| Mean corpuscular volume (MCV) | 91.43 (± 9.21) | 96.63 (±10.18) |
| Monocyte distribution width (MDW) | 24.43 (± 4.3) | 28.45 (±6.82) |
| MDW ≥20 | 5400 | 2450 |
| MDW < 20 | 1345 | 1034 |
| Mean platelet volume (MPV) | 8.54 (±0.92) | 8.74 (±1.05) |
| Red blood cell (RBC) count | 4.14 (±0.74) | 4.1 (±0.920) |
| Red cell distribution width (RCDW) | 16.67 (± 2.29) | 13.46 (±2.57) |
| White blood cell (WBC) count | 11.51(±5.35) | 13.43 (±6.41) |
| Platelet count | 218.71 (±113.32) | 249.23 (±114.11) |
| Eosinophil count | 0.13 (±0.12) | 0.11 (±0.10) |
| Lymphocytes count | 1.51 (± 0.61) | 1.37 (±0.72) |
| Monocyte count | 0.91 (± 0.34) | 0.76 (±0.43) |
| Neutrophil count | 9.53 (±5.31) | 10.51 (±5.23) |
| Basophil count | 0.18 (±0.) | 0.05 (±0.03) |

#### Data pre-processing, missing data, and feature engineering

The original input data consisted of seven files with comma separated values (CSV) that included more than 37 million distinct data elements. The original data files were processed using a data cleaning and feature engineering workflow developed by our group using Python Pandas. We implemented multiple pre-processing steps to address the issues of unbalanced data, missing values, and outlier values. All missing numeric features were imputed with mean, and median based on their distribution (listed in Table S1 in supplementary material), while outlier features were clipped within the 5th and 95th percentile values at the lower and upper end of the distribution, respectively. We used the *Standardscalar* package to normalize the raw data. The original data consisted of 66% patients with a diagnosis of suspected infection and 34% patients with severe sepsis-related outcomes. To address this imbalance between the number of patients in each category in the training data, we used a well-known method for data augmentation called Synthetic Minority Oversampling Technique (SMOTE) [30]. However, the dataset used to evaluate the trained ML models (test data) did not include any augmented data generated by the SMOTE library to prevent data leak.

We used the *imblearn* package (version 0.9) of the SMOTE library to generate additional records representing patients in the severe sepsis category, which resulted in a balanced dataset. We describe additional details of the SMOTE-based approach that was used to augment the dataset in Section 1.1 of the supplementary material. The training data set consisted of 10,792 patients (80% of the total data) with 50% (5396) with severe sepsis, and 50% (5396) patients with suspected infection. The dataset used for evaluating the trained ML algorithms consisted of 2046 patients (20% of the total data) with 34% (697) patients with severe sepsis and 66% (1349) with suspected infection (test dataset did not include any data generated by the SMOTE library).

### 2.2. Machine learning algorithms and interpretability methods

We used four ML algorithms in the study, namely logistic regression (LR), support vector machines (SVM), random forest (RF), and deep neural network (DNN). LR is a well-known ML algorithm that uses a logistic function as a classification boundary to assign probabilities to labels assigned to data element while minimizing the loss value on a data set. LR has been used as a statistical measure in quick Sequential Organ Failure Assessment (qSOFA) measure to identify patients at risk of sepsis using multiple clinical parameters [28]. SVMs are kernel-based classification algorithms that have been highly effective for binary classification tasks by balancing efficient training algorithms with the ability to represent complex functions. RF is the third classical machine learning model that uses multiple decision trees with specialization for a learning task and the result is based on the aggregation of the output of the decision trees. Finally, we implemented a DNN architecture using TensorFlow Keras API with the input and output layers consisting of 32 nodes and 1 node respectively. We describe the implementation details, parameter values, and architecture of the four ML algorithm in Section 1.2 of supplementary materials. The hyperparameter values for these ML algorithms were tuned using a Keras grid search library [31].

#### Interpretability of results generated by ML algorithms

Interpretability methods are used to characterize changes in the output of ML algorithms based on changes in their input values and provide insights into the role of individual input values in the performance of the ML algorithms [31]. In contrast, explainability methods are focused on the dynamics of ML algorithms instead of the characterizing the impact of input features on the results [31]. In this study, we use ML interpretability methods. We used two model agnostic interpretability methods, namely Shapley Additive Value (SHAP) [32] and Local Interpretable Model-Agnostic Explanation (LIME) [33] to interpret the results of ML algorithms with high accuracy.

Both the LIME and SHAP methods use linear models that approximate the original ML algorithms together with perturbations to the original input values to generate an explanation with lower complexity [32, 33]. The LIME model computes the importance of the input features by heuristically defining the complexity of the model used for explanation together with cosine value used as distance metric for perturbing the input values [33, 34]. The SHAP method uses the framework of cooperative game theory to define the complexity of explanation model and the distance measure used in perturbing the input values [32, 34]. Using these two interpretability models, we systematically evaluated the contribution of individual hematologic parameters, including MDW, and vital signs data. The results of our interpretability analysis are discussed in the next section.

## 3. Results

In the first phase of our two-phase analysis workflow, we defined seven data models based on clinical input from co-authors YT, SA, and DK, who used the availability of relevant data throughout the clinical evaluation process to guide their decision. New data elements were added incrementally to each model to systematically characterize individual contributions of a data element, which resulted in the following set of data models:

- **Data Model 1**: The baseline model consists of CBC standard data elements (hematocrit, hemoglobin, MCH, MCHC, MCV, MPV, platelet count, RBC count, RDWC, and WBC count).
- **Data Model 2**: This model adds laboratory results for CBC differential test to the baseline model (additional data elements were: eosinophil count, lymphocytes count, monocyte count, neutrophil count, and basophil count).
- **Data Model 3**: MDW was added to data model 2 to create data model 3.
- **Data Model 4**: This model adds vital signs data (respiratory rate, systolic and diastolic blood pressure, and temperature) to data model 3.
- **Data Model 5**: This model removes MDW as an input data element, which enables a characterization of the impact of other hematologic parameters and vital signs on the accuracy of ML algorithms.

In addition, we created two additional data models, that is, data model 6 with only MDW as a feature and data model 7 with both MDW and WBC count as features. Data model 7 corresponds to the model used in the study by Crouser et al., which was submitted to the US FDA as part of the approval process for MDW as an independent biomarker for sepsis [13].

### 3.1. Classification of patients using ML algorithms

The ML algorithms performed a binary classification task with each patient classified into either severe sepsis-related outcome category or suspected infection category. All the four ML algorithms were implemented over the seven data models. The ML algorithms were evaluated using the test dataset and the performance of the algorithms is reported using the area under the receiver operating characteristic (ROC) curve (AUC), and the error matrix consisted of true positive, false positive, true negative, and false negative values. We report the AUC value as the aggregate of true positive rate (TPR) and false positive rate (FPR) with TPR defined as the ratio of true positive and positive while FPR is defined as 1-true negative rate (ratio of true negative and negative) [35]. Table 2 lists the performance of the four ML algorithms in terms of the AUC (with confidence interval) over the first five data models and Table S2 in the supplementary material lists the performance of ML algorithms over data models 6 and 7.

**Table 2:**
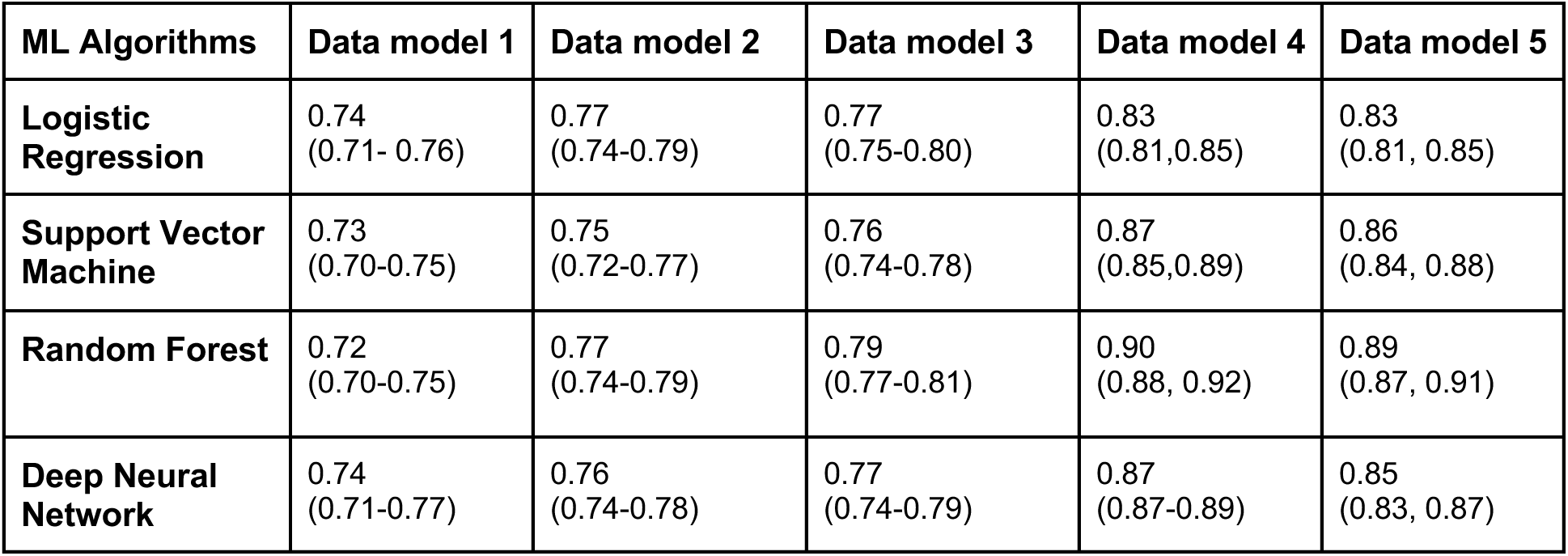
Performance of the four ML algorithms over the five data models.

The DNN and LR ML algorithms have the highest accuracy among the four ML algorithms for data model 1 with an AUC of 0.74. The accuracy values of SVM, and RF were 0.73, and 0.72 respectively. Supplementary Figure S2(a) shows the ROC plot of the performance of the four ML models over data model 1. As additional data elements from CBC differential test were added in data model 2, the performance of all the four ML algorithms improves with the highest improvement seen in the RF model (AUC = 0.77); the RF and LR algorithm still outperforms the other two algorithms with an AUC of 0.77. Supplementary Figure S2(b) shows the ROC plot of the performance of the four ML models for data model 2.

Figure 2 shows the comparative performance of the four ML algorithms as MDW measure is added in data model 3 and followed by the addition of vital signs in data model 4. The addition of the MDW value in data model 3 (Figure 2(a)) marginally improves the performance of the three classical ML algorithms, namely LR with AUC = 0.77, SVM with AUC = 0.76, and RF with AUC = 0.86, while the DNN model shows greater improvement in the performance with AUC = 0.78 and AUC = 0.76 respectively. However, the addition of vital signs data (breathing rate, blood pressure, body temperature) in data model 4 leads to marked improvement in the performance of all the ML algorithms as compared to the addition of MDW (Figure 2(b)) with RF, DNN, and SVM performing considerably well with AUC of 0.90, 0.87, and and 0.87 respectively. The performance of the LR algorithm also improves with an AUC of 0.83. The role of vital signs data in detection of sepsis is well documented in previous studies and these results for data model 4 confirm the findings of previous studies. The results for data model 5 (Figure 2(c)) show that there is marginal decrease in performance of all four ML algorithms when MDW is removed as an input feature.

**Figure 2:**
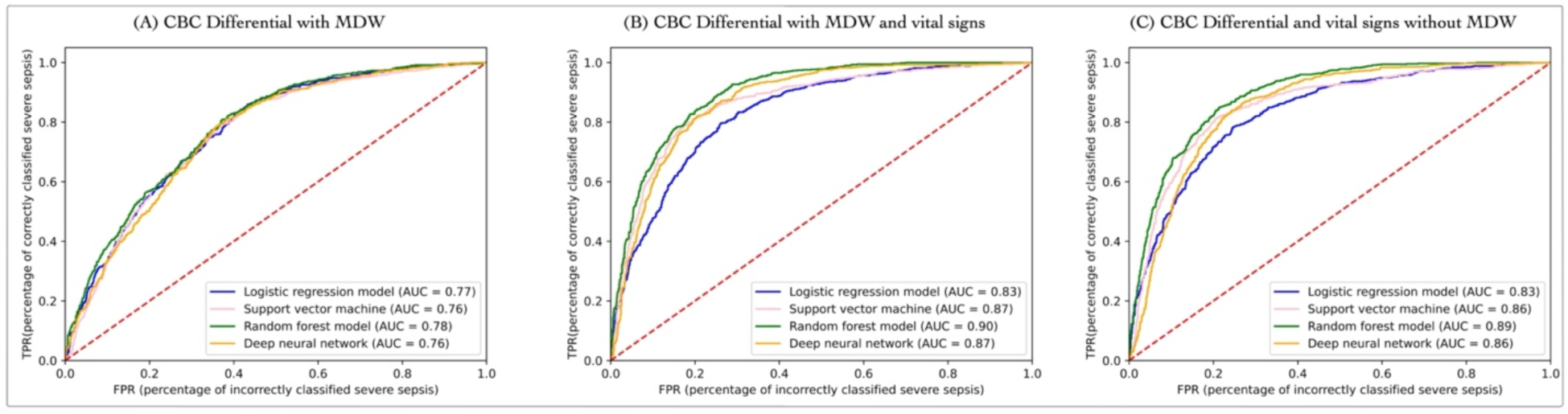
A comparative evaluation of the performance of the four ML algorithms in the detection of severe sepsis with and without the MDW value across three data models. (A) There is only marginal increase in the performance of the four ML algorithms with MDW and other hematologic test data (CBC standard and CDC differential). (B) The addition of vital signs data significantly improves the performance of all four ML algorithms. (C) In data model 5, the performance of the ML algorithms is only marginally affected by the removal of MDW as a feature, which shows that the contribution of MDW in detection of severe sepsis is attenuated in the presence of other hematologic parameters and vital sign data.

To quantify the change in accuracy values of the ML algorithms with and without MDW values in data model 3, data model 4, and data model 5, we performed a corrected t-test based on random subsampling test over the accuracy values reported by the ML algorithms [36]. The addition of vital signs in data model 4 led to a statistically significant improvement in the performance of all four ML models and the decrease in accuracy value reported for data model 5 was not statistically significant. This suggests that MDW plays a marginal role in screening patients with suspected infection for outcomes associated with severe sepsis and its relative importance is low as compared to other hematologic parameters and vital signs. We provide additional details of the corrected t-test in Section 1.3 of the supplementary material. In supplementary material Section 2, Figure S3 shows the results of the four ML algorithms for data model 6, which includes only MDW values for patient classification. Further, Figure S4 shows the results of the four ML algorithms for data model 7 that includes both MDW and white blood cell (WBC) count, which was used by the FDA for approval of MDW [13].

### 3.2. Interpretability of results generated by ML algorithms

We applied the LIME and SHAP interpretability methods to the RF and DNN ML algorithms with the highest accuracy values (AUC of 0.90 and 0.87 respectively) over data model 4 (with MDW as an input feature) to characterize the importance of individual hematologic parameters and vital sign as input features in screening for severe sepsis.

#### Relative importance of features for individual patients and for the study cohort

In the first step, two patients were randomly selected, that is, a patient with a severe sepsis-related outcome and another patient with infection. The two interpretability methods were applied to the classification result generated by the RF ML algorithm for these two patients. We also applied the SHAP method to the classification result generated for the two patients by the RF ML algorithm and used a decision plot to visualize the feature importance scores (Figure 3(A)). The list of features in Figure 3(A) are ordered by the descending order of feature importance scores. As the SHAP values represent the difference between the expected and actual outputs of a ML algorithm, the decision plots for the two patients start at the base of the y-axis, which corresponds to the expected value of the result generated by a ML algorithm. In the next step, SHAP value for each feature is progressively added to the expected value leading to the final classification outcome generated by the ML algorithm [32]. We see that for both the patients, respiratory rate, age, monocyte count, among other hematologic biomarkers contribute significantly to the final classification result. However, MDW value has only a marginal contribution to the final output of the RF algorithm for both of the patients.

**Figure 3.**
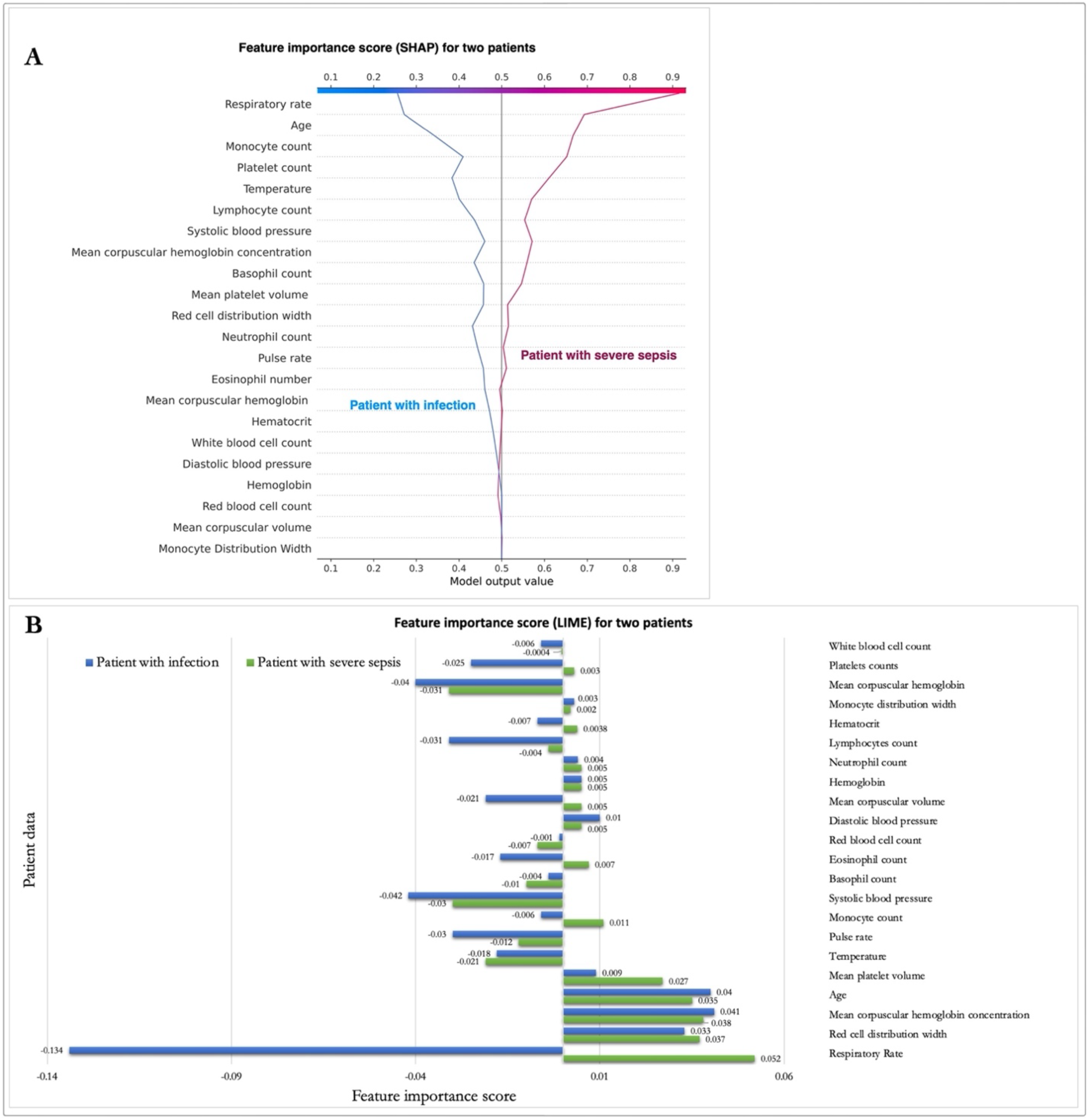
Feature importance scores of data elements for two randomly selected patients (one with severe sepsis and another with suspected infection) using LIME and SHAP interpretability methods with a decision plot (A) and a waterfall plot (B). The feature importance scores are computed for the RF ML algorithm.

The waterfall plot for the two patients using the LIME method (Figure 3(B)) shows that vital signs, and hematologic biomarkers have high feature importance scores, that is, they have a significant contribution to the final classification result of the RF ML algorithm. For the patient with infection, hematologic data elements (e.g., RCDW, MCHC, and mean platelet count) contribute significantly to the classification output. In contrast, the LIME method assigned high feature importance score to vital signs, including respiratory rate, and age, together with hematologic parameters, such as RCDW, MCHC, and monocyte count for the patient with infection. Although there are several common data elements that have high feature importance score computed by the LIME method for both the patients (e.g., RCDW, and MCHC), the feature importance score of MDW is low for both the patients.

Building on the interpretability results for the two randomly selected patients, we computed the feature importance scores for all the patients in the study cohort that forms the test dataset (n = 2046) (Figure 4). The pattern of feature importance scores for the study cohort are similar to the results generated for individual patients with vital signs data, and multiple hematologic parameters and vital signs contributing significantly to the classification output generated by the RF ML algorithm. However, MDW continues to have a relatively low feature importance scores as compared to other hematologic parameters and vital signs. Specifically, the feature importance scores for MDW are 0.021 and 0.003 as computed by SHAP and LIME methods respectively. In contrast, the RCDW has feature importance scores of 0.1 (SHAP) and 0.06 (LIME); MCHC has scores of 0.01 (SHAP) and 0.006 (LIME); and lymphocyte count has scores of 0.05 (SHAP) and 0.023 (LIME). These cohort-level results validate the current clinical practice of relying on vital signs together with readily available CBC standard or CDC differential values for screening ED patients for severe sepsis related-outcomes. Figure S5 in the supplementary material visualizes the feature importance scores of data elements computed using LIME and SHAP methods using the DNN ML algorithm.

**Figure 4.**
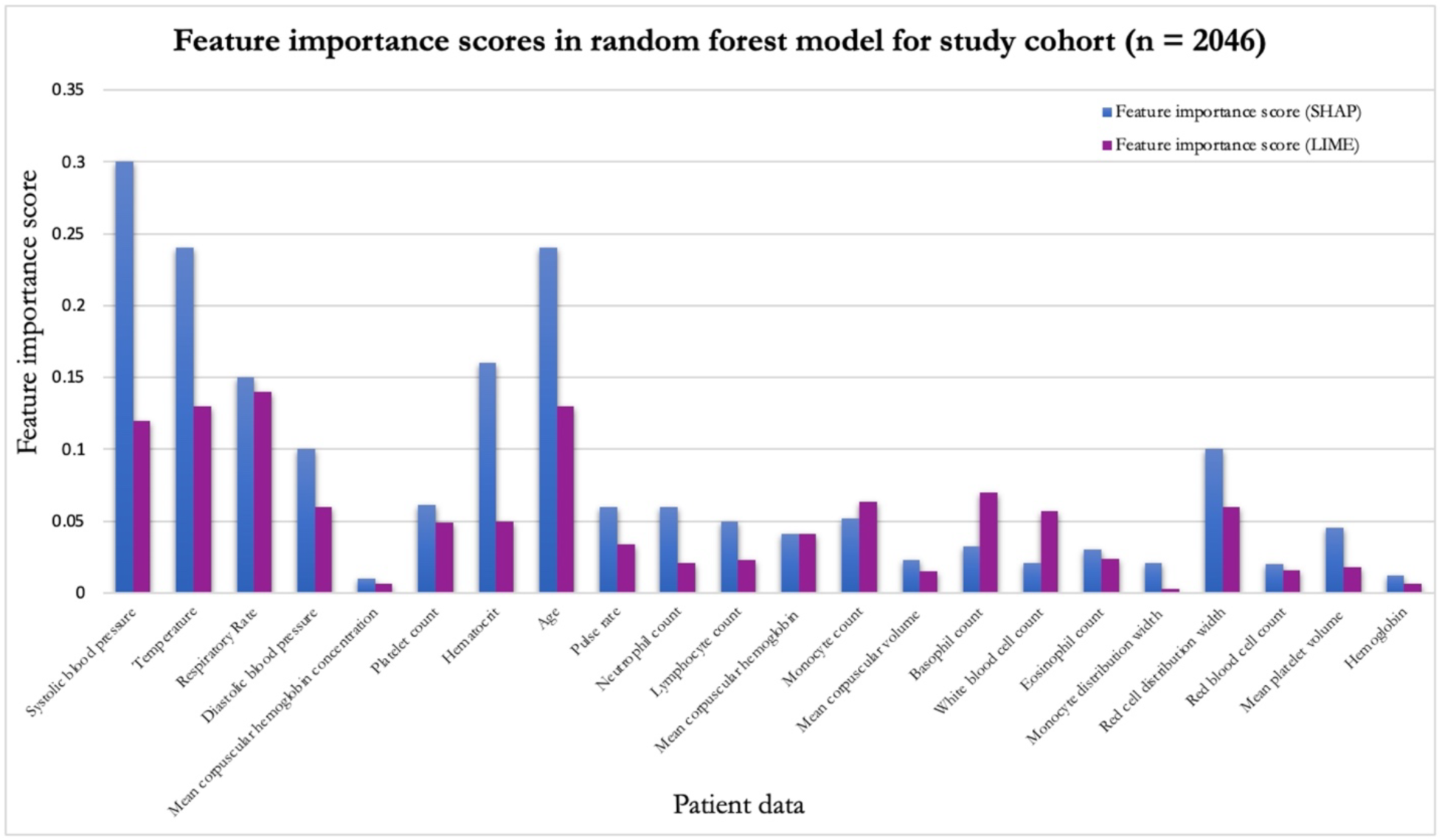
The feature importance scores calculated using SHAP and LIME methods for all the patients in the study cohort used as test dataset with the RF ML algorithm.

### 3.3. Impact of variation in values of individual features on results of the ML algorithm

We selected five features with the highest feature importance scores together with MDW values and plotted the dependence plots to illustrate how variation in values of these features impact the classification output of the RF algorithm. Figure 5 shows that there is a clear increase in the risk of severe sepsis in patients if the respiratory rate increases above 20 breaths per minute. Similarly, patients with low systolic blood pressure are at an increased risk of severe sepsis. The finding that changes in blood pressure value is also associated with the risk of severe sepsis is consistent with current clinical knowledge as these changes in blood pressure herald the development of septic shock.

**Figure 5.**
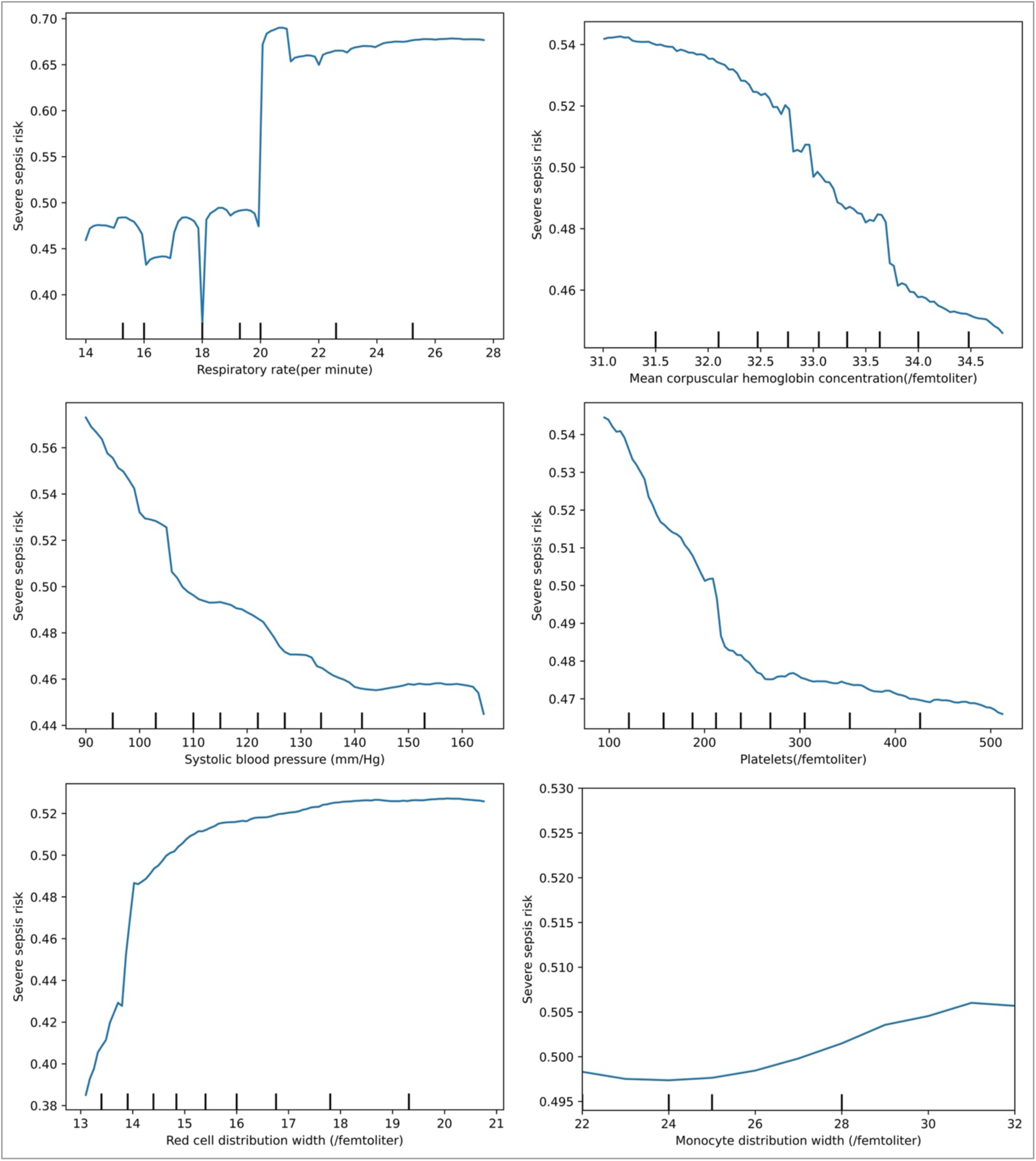
The partial dependence plots of the five features with the highest feature importance scores and MDW show the impact of changes in values of these features on screening for severe sepsis. These values are computed using the patient records in test dataset for the output generated by the RF ML algorithm.

The association between hematologic parameters such as platelet count, MCHC, and MDW with risk of severe sepsis is also highlighted by these dependence plots. These data show that as MDW value increases over 23, there is increased risk of severe sepsis-related outcome; however, the rate of increase is gradual with the risk value increasing from 0.506 to approximately 0.510. These finding correspond to results from previous studies, such as a study by Hou et al., in pediatric emergency department, which found that MDW value greater that 23 is associated with pediatric sepsis [37]. There is well-known association between RCDW and platelet count in severe sepsis and the partial dependence plots highlight these association with an exponential increase in severe sepsis risk as the RCDW score increases above 13 units. A decrease in platelet count increases the risk of severe sepsis and as the platelet count decreases below 200,000 per microliter there is a dramatic increase in risk of severe sepsis.

#### Interaction effect between MDW and other data elements

Though the feature importance of MDW in the collective model was modest, we explored the interaction effect of data elements with high feature importance scores and MDW using scatter dependence plots (Figure 6). The scatter dependence plot of respiratory rate value and the SHAP score of respiratory rates with respect to MDW values (Figure 6(a)) show distinct vertical patterns of coloring at specific values of respiratory rate. These vertical patterns correspond to several patients who were classified by the RF ML algorithm with equal respiratory rates but varying MDW values, which suggests that MDW value does add some prognostic value to respiratory rate due to interaction effect; however, the interaction effect pattern is not robust. A similar pattern, which is not robust, is noted in the scatter plot for systolic blood pressure. Either of these patterns may be due to the added discriminative value of MDW value together with other manually ascertained data that are subject to high inter-observer variability and rounding [38]. Nonetheless, these patterns do not demonstrate a prognostically valuable interactions effect of MDW with other hematologic parameters and vital signs with high feature importance scores.

**Figure 6:**
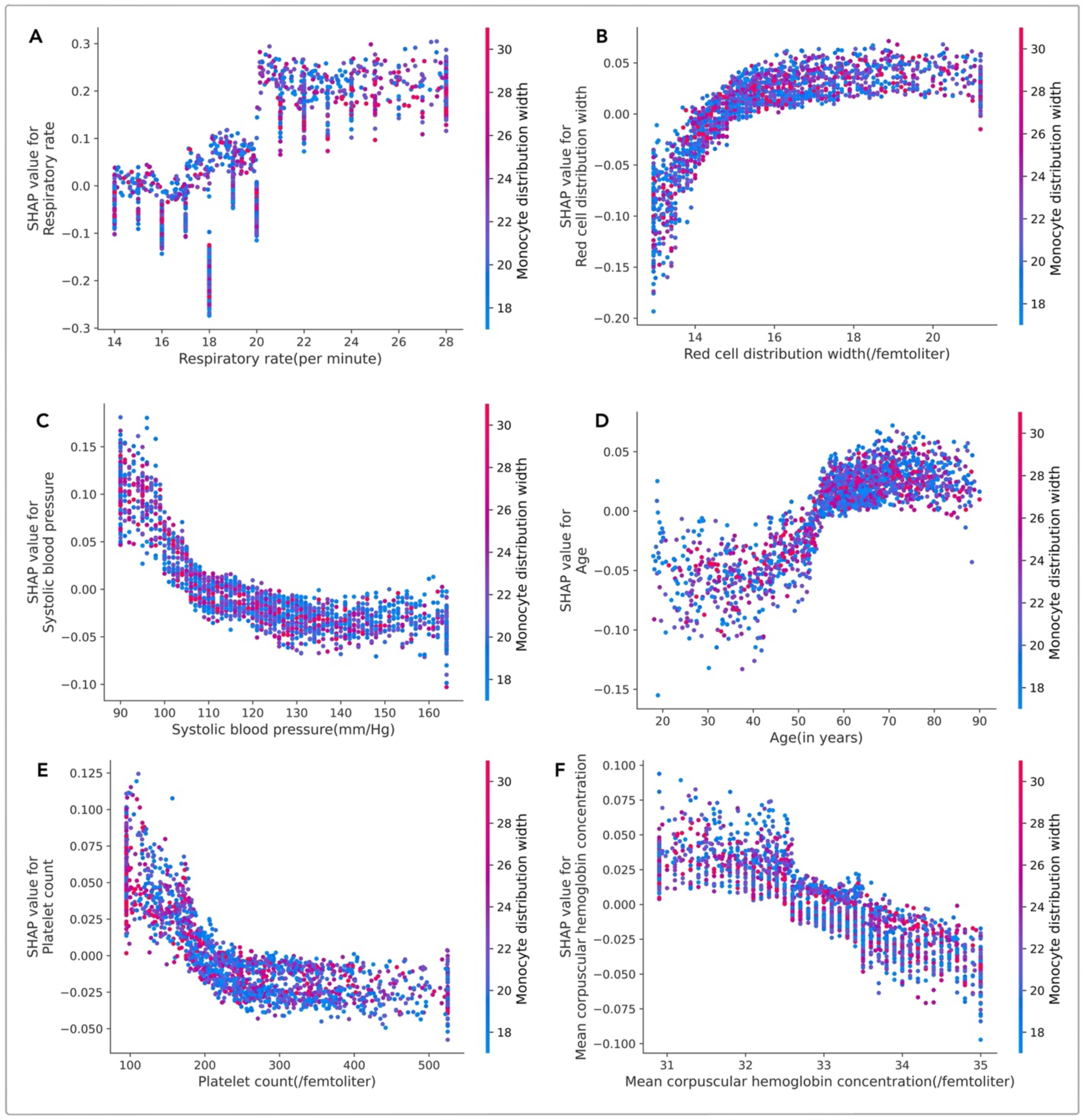
The interaction effect between MDW and input features with high feature importance scores generated by the RF ML algorithm shows that MDW has some interaction effect with vital signs readings (respiratory rate and blood pressure), age, and platelet count (these scatter dependence plots use SHAP values).

## 4. Discussion and Conclusion

This study combines an ensemble of high accuracy ML models with interpretability methods to characterize the relative importance of MDW with respect to other hematologic parameters and vital signs to screen for patients with severe sepsis-related outcomes.

### 4.1. Clinical significance and limitations

Our results demonstrate the value of an ensemble approach in identifying the additional prognostic value of MDW as a relatively novel hematologic biomarker in the detection of severe sepsis. While MDW has been shown to independently predict the risk of sepsis and more severe septic phenotypes; however, this study shows that when other routine laboratory test results and vital signs data are considered, the contribution of MDW in detecting severe sepsis is grossly attenuated. We note that these results do not diminish the relevance or importance of the physiologic phenomena represented by the MDW value, and it does not negate its value in other potential use cases. For example, the study by Hossain et al., found that MDW can add useful prognostic information with regards the prediction of hypoxemic respiratory failure early in patients with COVID-19 [20].

A key attribute of this study is that the analysis uses a large cohort of patients with hematology parameters drawn and analyzed through routine care processes. The patient population used in this study is diverse, thereby increasing the potential for generalizability of these results across different healthcare settings. However, we note that the conclusions of these study are limited to a single site study. Further, although we leveraged the outcome measures used by Sepsis-3 and qSOFA, that is, infection with a three-day ICU stay or death; however, the study lacks the use of a gold standard diagnosis for severe sepsis. Thresholds for ICU admission and length of stay are also likely variable, making it important that our findings be externally validated in different settings. Additionally, our analysis hinges on the availability of data only in patients who had a CBC with differential panel ordered in the first 24 hours. These patients likely differ from those that do not have cell differential data by virtue of presentation or critical illness. By design, this limits our conclusions for the entirety of the patient population presenting to the ED. This can only be done in a study if the cell count with differential data are obtained for all patients presenting to the ED, which is an expensive but important proposition to fully evaluate the return on investment of biomarkers like MDW in this setting.

Nonetheless, the results of this study should provide caution for the adoption of a single biomarker for sepsis prognostication and provides a roadmap for developing a robust evaluation of the significance of individual parameters as a marker with the availability of potentially more prognostic, pre-existing data.

We note that the application impact of the ML methods developed in this study can be enhanced if healthcare systems can overcome present day barriers to adopt advanced analytics and implement them at the point of care [39]. In the absence of a pragmatic framework to leverage ML workflow, existing healthcare systems may be forced to rely on individual prognosticators like MDW or rule-based scoring systems instead of more advanced clinical decision support systems. Recent encouraging developments in sepsis care have highlighted growing evidence that prediction-based algorithms have clear clinical benefits on patient outcomes, especially in the context of the risk due to severe sepsis [4, 6].

### 4.2. Post-hoc interpretability methods for ML algorithms and their limitations

The two interpretability methods used in this study are part of a larger group of post-hoc methods used for providing insights into results generated by ML algorithms. Both LIME and SHAP methods build simpler models that represent the original ML algorithms and use input values that are representative of the original input values to generate results that closely match the original results. The representative models that are trained in both the methods are more interpretable as compared to the original ML model; therefore, they are used to measure the contribution of individual input features on the results.

Recent work has highlighted some of the limitations of these post-hoc interpretability approaches as they can be modified by an adversarial model that can distinguish between input features provided by interpretability models, such as LIME and SHAP, and the original input values [34]. Although, this is an important limitation of post-hoc interpretation methods, we note that this adversarial approach is viable in scenarios where a third party provides the interpretability services. In this study, we implemented the interpretability models without the use of any third party trained representative model or scaffolding, which limited the risk of unverified modification of feature importance scores in this study.

In conclusion, the study demonstrated that the relative importance of MDW as a biomarker for prognostication of sepsis-related outcomes is limited and the method developed in this study using an ensemble of high accuracy ML algorithms together with model agnostic interpretability methods is a generalizable approach for informed clinical decision making in healthcare systems.

## Data Availability

The machine learning workflows and performance metrics were implemented using the Scikit libraries. The individual patient records cannot be made publicly available due to regulatory reasons. Models and data can be made available on request; however, this requires the execution of a data transfer agreement approved by the participating institutions together with an Institutional Review Board (IRB) or equivalent ethics approval for the proposed study.

## Contributors

DPU, YT, SSS conceived and designed the study. DPU, YT, SSS participated in the extraction of EHR reports, selection of features, and curation of the reports. DPU and SSS performed feature engineering and machine learning with interpretability. KP performed the statistical analysis. SA and DK contributed to the creation of the study cohort and the use of the ED workflow. DPU, YT, SA, DK, and SSS reviewed the clinical data. DPU, YT, SA, and SSS wrote the initial draft of the manuscript. All authors subsequently read, revised, and approved the final version. All authors had access to all the data in the study and DPU, YT, KT, and SSS verified the data and had final responsibility to submit for publication.

## Acknowledgements

This research was funded in part by the grants from the US National Institutes of Health (NIH): U24EB029005, R01DA053028, the US Department of Defense (DoD) grant W81XWH2110859, and the Clinical and Translational Science Collaborative of Cleveland, which is funded by the NIH, National Center for Advancing Translational Sciences, Clinical and Translational Science Award grant, UL1TR002548. The content is solely the responsibility of the authors and does not necessarily represent the official views of the NIH.

## Supplementary Material

### 1. Supplementary Methods

We discuss additional details of the method used in this study to characterize the relative importance of hematologic parameters, including monocyte distribution width (MDW), and vital signs in prognosticating patients with suspected infection.

#### 1.1 SMOTE algorithm for data augmentation

As compared to traditional oversampling and under sampling methods used for addressing the challenge of learning over imbalanced data, the SMOTE algorithm uses randomized interpolation of features from a set of instances representing the minority classes to generate new instances of minority classes [40]. The SMOTE algorithm has been shown to be effective in a variety of applications and it addresses the limitation of oversampling techniques for overlapping or disjunct data and under sampling that increases the variance of classifiers and potentially leaves out important instances of majority classes during training [41].

The SMOTE algorithm does not generate new variations in the model and uses randomization to generate new records using the K-nearest methods from the feature space. We applied K nearest methods to the input data where K is selected as 5. There are different sampling strategies available in the SMOTE library, such as “minority”, “not minority”, “not majority”, “all”, and “auto”, among others. In this study, the SMOTE library oversampled records meeting the primary clinical outcome to generate additional records of patients with outcomes attributable to sepsis. We used auto as a sampling strategy which resamples all classes. The resulting dataset included 6745 records of patients with suspected infections and 6745 records of patients meeting the primary clinical outcomes.

#### 1.2 Machine learning algorithms and ML interpretability methods

Logistic regression (LR) is a well-known machine learning (ML) algorithm that uses a logistic function as a classification boundary to assign probabilities to labels assigned to data element while minimizing the loss on a data set. LR has been used as a statistical measure in quick Sequential Organ Failure Assessment (qSOFA) measure to identify patients at risk of sepsis using multiple clinical parameters [28]. Support Vector Machines (SVM) are kernel-based classification algorithms that have been highly effective for binary classification tasks by balancing efficient training algorithms with the ability to represent complex functions. Random forest (RF) is the third classical machine learning model used in the study that uses large number of decision trees with specialization for a learning task and the result is based on the aggregation of the output of the decision trees. Finally, we implemented a Deep Neural Network (DNN) architecture using TensorFlow Keras API with the input and output layers consisting of 32 nodes and 1 node respectively. We implemented the LR classifier from the scikit-learn package with inverse of regularization (*C=1.0*), solver (*lbfgs*), tolerance for stopping criteria (*tol = 0.0001*), and *l2* as a penalty. We implemented the SVC classifier of the support vector machine (SVM) model in the scikit-learn package with regularization, *C = 10*, kernel = *poly* with degree of *kernel function = 4*, and probability estimates set as *true*. The random forest (RF) model in the scikit-learn package was used in the study with 700 trees in the forest, the maximum depth of the tree depth was set as 25 after training and validation phases. The deep neural network (DNN) architecture also included 4 hidden layers with a variable number of nodes with rectified linear unit (ReLU) activation function used for each hidden layer. The model used Adam optimizer with Binary_crossentropy as the loss function and learning rate of 0.001. The final output layer used a sigmoid activation function to generate a probability of class membership value between 0 and 1.

Both the Shapley Additive Value (SHAP) and Local Interpretable Model-Agnostic Explanation (LIME) methods use linear models that approximate the original ML algorithms together with perturbations to the original input values to generate an explanation with lower complexity [32, 33]. The LIME model computes the importance of the input features by heuristically defining the complexity of the model used for explanation together with cosine value used as distance metric for perturbing the input values [33, 34]. The SHAP method uses the framework of cooperative game theory to define the complexity of explanation model and the distance measure used in perturbing the input values [32, 34]. Using these two interpretability models, we systematically

#### 1.3 Statistical Analysis

We performed a corrected t-test based on random subsampling of our data to evaluate the relative importance of MDW and vital signs data in the classification of patients [36]. We performed five repetitions of random subsampling of an 80/20 data partition. For each repetition, we calculated the model accuracy of each of our algorithms (LR, SVM, RF, and DNN) for each data model (model 3, model 4, and model 5), and compared the results using a corrected resampled pairwise t-test. The test statistic can be calculated using the following formula, where:

1. n is the number of repetitions of random partitioning
2. a_i_ refers to accuracy value for a model A algorithm using the i^th^ random subset.
3. b_i_ refers to accuracy value for a model B algorithm using the i^th^ random subset.
4. σ^2^ estimated the variance of the n differences in accuracy between model A and model B.
5. n_1_ refers to the number of instances used for training, and
6. n_2_ refers to the number of instances used for testing.

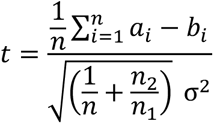

The p-values were calculated from the test statistic according to a t-distribution with 5 degrees of freedom (df = n-1). All calculations were performed in Python (version 3.10).

The results (with p = 0.05) show that the improvement in accuracy from model 3 to model 4 is statistically significant for all ML algorithms except the RF ML algorithm, which was approaching significance with a p value of 0.09. The change in accuracy between model 4 and model 5 is not significantly different. Model 4 and model 5 consistently differed in accuracy for DNN by a value of 0.01 for all 6 random partitions, thus due to a lack of variation, statistical comparison could not be completed for the DNN algorithm.

### 2. Tables and Figures

**Table S1:** List of data elements with missing or out of range values.

| Features | Data type | Missing or out of range value, n (%) |
| --- | --- | --- |
| Basophil count | Float | 735 (7.19%) |
| Eosinophil count | Float | 735 (7.19%) |
| Lymphocyte count | Float | 735 (7.19%) |
| Monocytes count | Float | 735 (7.19%) |
| Neutrophil count | Float | 735 (7.19%) |

**Table S2:** Performance of the four ML algorithms over data model 6 and data model 7.

| ML Algorithms | Data Model 6 | Data Model 7 |
| --- | --- | --- |
| **Logistic Regression** | 0.54 (0.51- 0.56) | 0.64 (0.61-0.67) |
| **Support Vector Machine** | 0.56 (0.53-0.58) | 0.65 (0.62-0.67) |
| **Random Forest** | 0.59 (0.56-0.61) | 0.64 (0.62-0.67) |
| **Deep Neural Network** | 0.58 (0.56-0.61) | 0.67 (0.64-0.69) |

**Figure S7:**
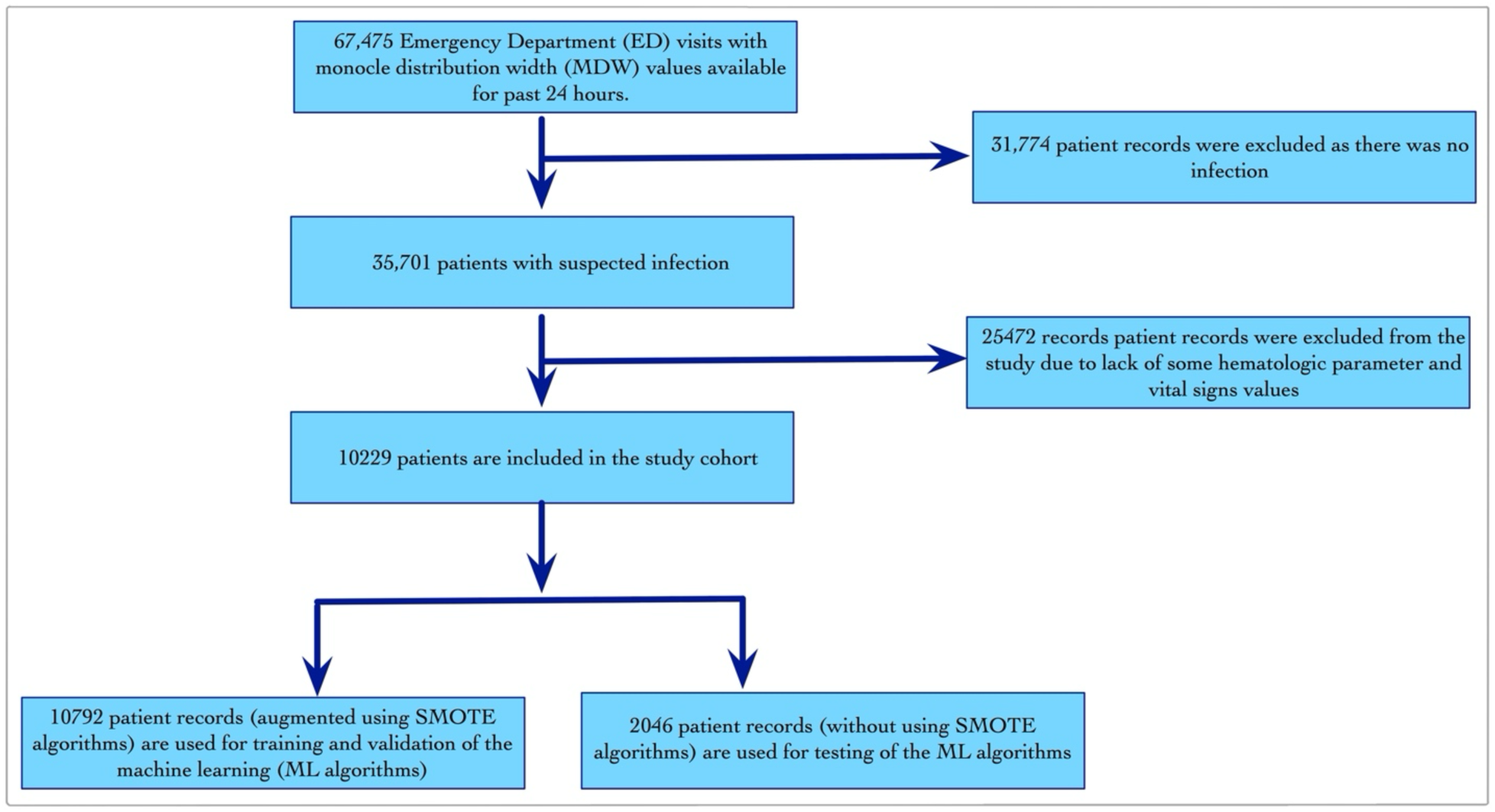
The inclusion exclusion criteria applied to all ED visits at the MetroHealth hospital system between January 1st, 2019, and December 24th, 2023, to create a cohort of 10,229 patients. The records in the study cohort were augmented using the SMOTE algorithms to address the imbalance between patients with suspected infection and those with suspected infection and the primary clinical outcome in training dataset.

**Figure S8:**
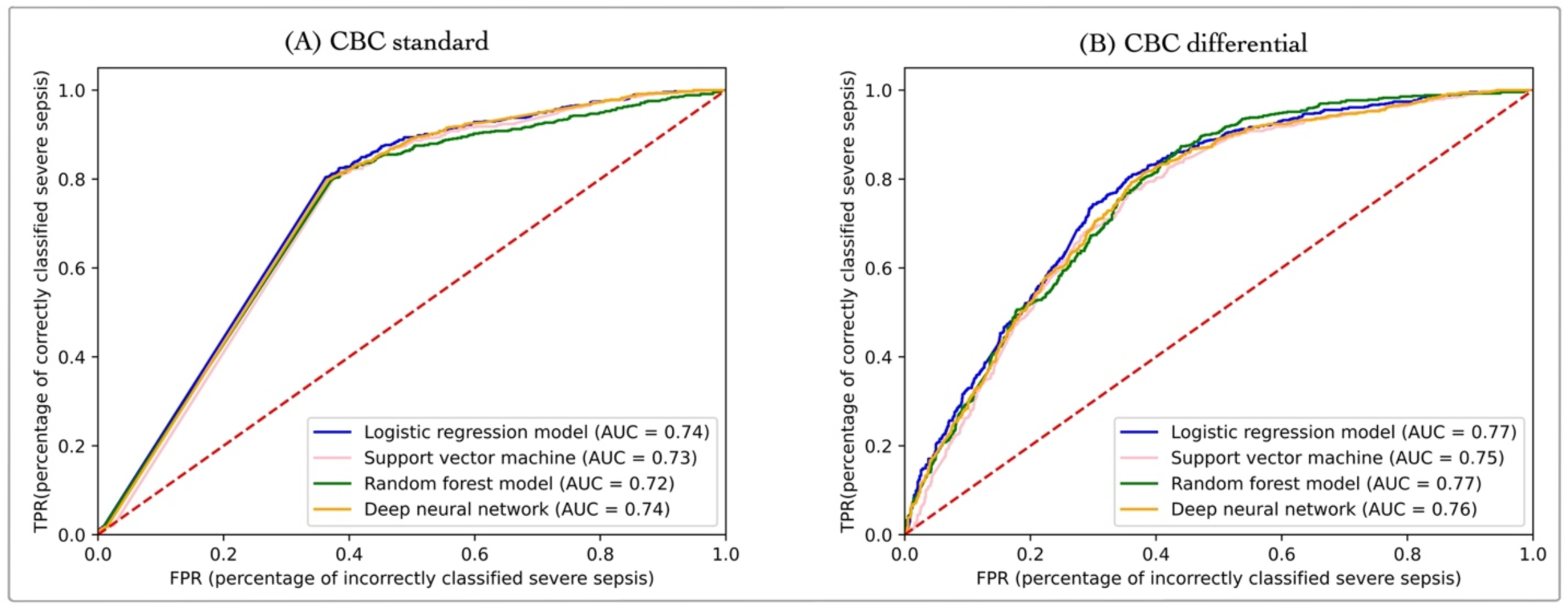
A comparative evaluation of the four ML algorithms over data model 1 ((A) CBC standard data elements) and data model 2 ((B) CBC differential data elements). The addition of CBC differential data elements improves the performance of the ML algorithms.

**Figure S9:**
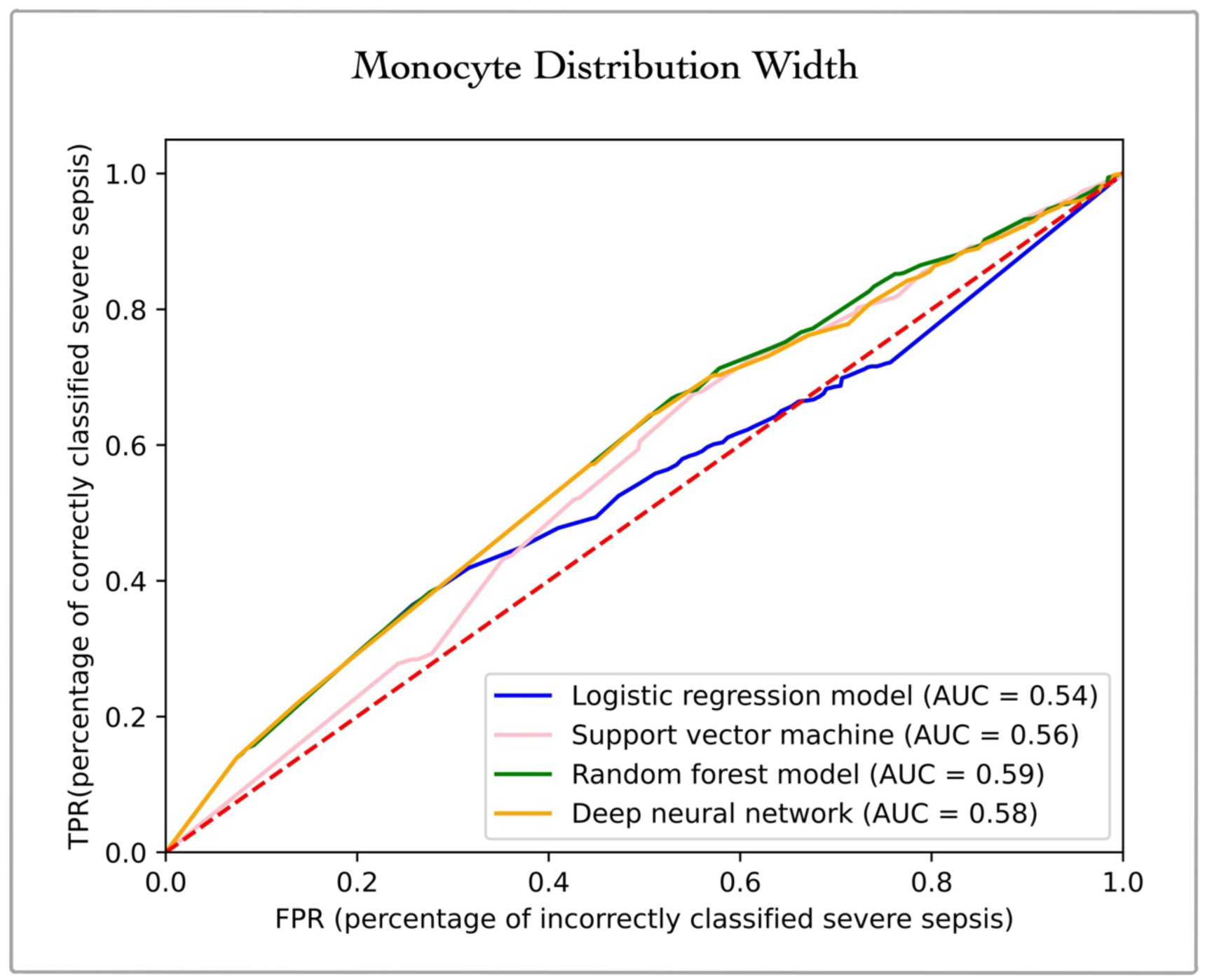
The performance of the four ML algorithms in data model 6 with only MDW values shows low accuracy values (slightly better than random chance) with the highest value reported for RF (AUC = 0.59).

**Figure S10:**
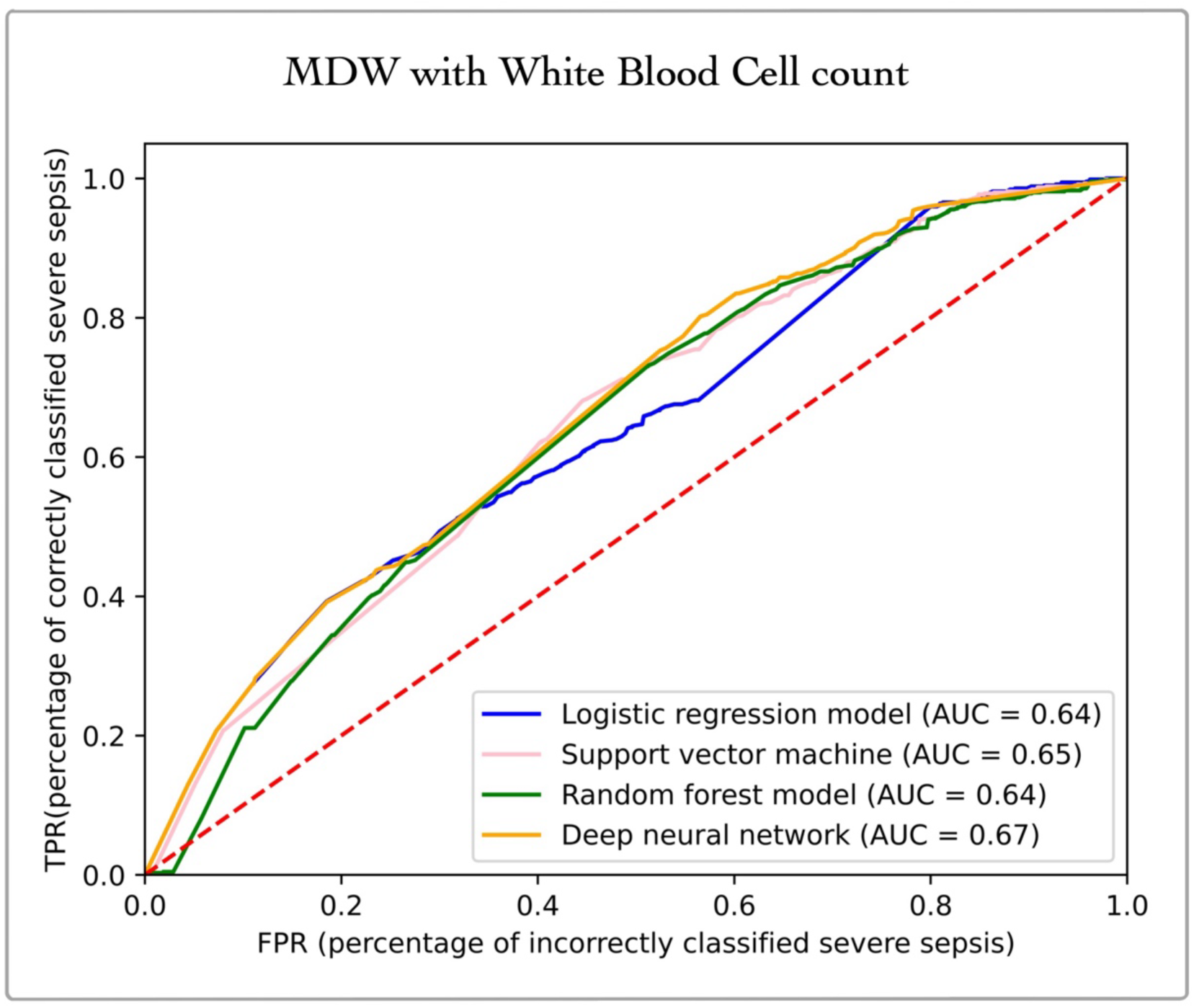
Performance of the four ML algorithms in data model 7 (with MDW and white blood cell count) shows low accuracy values with the highest value reported for DNN (AUC = 0.67).

**Figure S11:**
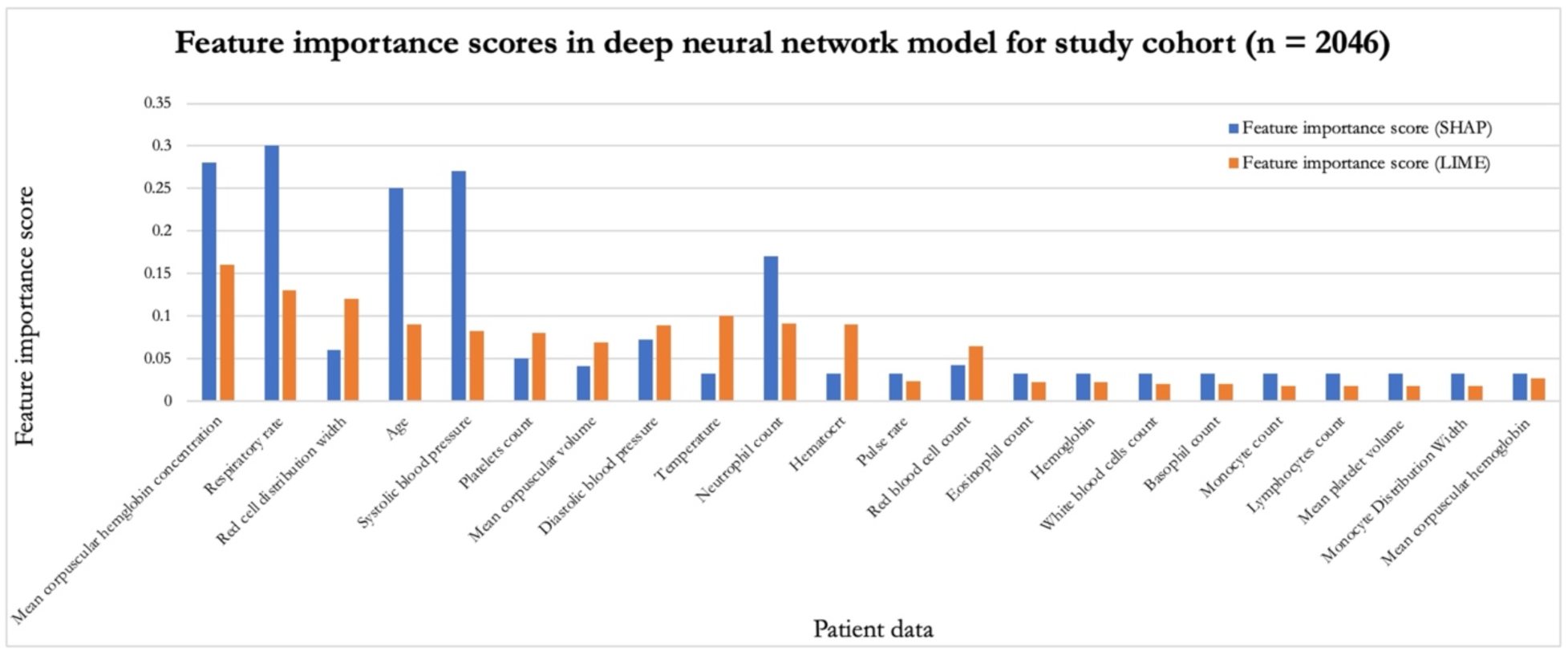
The feature importance scores computed using SHAP and LIME methods for all patients in the study cohort using deep neural network (DNN).

